# Impact of the COVID-19 pandemic on Suicide and Self Harm among Patients Presenting to the Emergency Department of a Teaching Hospital in Nepal

**DOI:** 10.1101/2020.10.16.20213769

**Authors:** Roshana Shrestha, Shisir Siwakoti, Saumya Singh, Anmol Purna Shrestha

## Abstract

**Background:** The COVID-19 pandemic is a global challenge that is not just limited to the physical consequences but also a significant degree of a mental health crisis. Self-harm (SH) and suicide are its extreme effects. The aim of this study was to provide an overview of the impact of the COVID-19 pandemic on the occurrence and clinical profile of suicide and SH in our ED.

**Methods:** This is a cross-sectional observational study conducted in the ED of a tertiary care center. Records of all fatal and nonfatal SH patients presenting to the ED during the lockdown period (March 24-June 23, 2020; Period1), matching periods in the previous year (March 24-June 23,2019; Period 2) and 3 months period prior (December 24 2019-March 23, 2020; Period 3) was included by searching the electronic medical record (EMR) system. The prevalence and the clinical profile of the patients were compared between these three periods.

**Results:** A total of 125 (periods 1=55, 2=38, and 3=32) suicide and SH cases were analyzed. The cases of suicide/SH had increased by 44% and 71.9% during the lockdown period in comparison to the period 2 and 3. Organophosphate poisoning was the most common mode. Females were predominant in all three periods with a mean age of 32 (95%CI: 29.3-34.7). There was a significant delay in arrival of the patients in period 1 (p-value=0.045) with increased hospital admission (p-value =0.009) and in-hospital mortality (18.2% vs 2.6 % and 3.1%) (p-value=.001).

**Conclusion:** We found an increase in patients presenting with suicide and SH in our ED during the pandemic which is likely to reflect an increased prevalence of mental illness in the community. We hope that the result will prime all mental health care stakeholders to initiate mental health screening and intervention for the vulnerable population during this period of crisis.

## INTRODUCTION

The COVID-19 pandemic is presenting a global challenge not just in terms of infectious disease but also for mental health. The pandemic rapidly disseminated across the world with the first reported case in Nepal on January 25, 2020 and the first reported death on May 14, 2020.[1] Nepal responded with a nationwide lockdown from March 24, 2020. The increasing number of infections and uncertainty induced a substantial fear and concern leading to stress and anxiety which was superimposed by lockdown restrictions, financial breakdown, lack of physical contact with other family members and friends.[2] The consequences of pandemic and lockdown on socioeconomic, mental health, and other aspects of Nepalese society are immense.[3] These alarming conditions may exacerbate the suicidal rate which is already high in our part of the world.

Suicide and self-harm (SH) are a serious public health problem; however, it is preventable with timely, evidence-based, and often low-cost interventions. Every year approximately 800 000 people commit suicide and many more attempt it. In 2016, it was listed as the second leading cause of death among 15-29 year-olds worldwide.[4] Suicide is already a key public health concern in Nepal and was ranked 7th by suicide rate globally by 2014. The World Health Organization (WHO) reports an estimated 6,840 suicides annually or 24.9 suicides per 100,000 people in our country.[5] In addition, civil conflict and the 2015 earthquake have had significant contributory effects.[6,7]

Recent studies conducted during the COVID-19 pandemic have revealed high levels of stress, anxiety, and depression in the community.[8–10] Previous studies conducted after the epidemic outbreak of Severe Acute Respiratory Syndrome (SARS) in 2003 revealed a significant increase in the elderly suicidal rate in Hong Kong.[11] Few publications reporting COVID-19 related suicide are published.[12–16] Increased cases of suicide have been reported in police stations all over Nepal since the lockdown period.[17] However, we found no publications in regards to COVID-19 related suicides presenting to the ED in developing countries. Most cases of SH present to the ED, therefore this study is the first of its kind done in an acute care setting to address this crucial issue related to mental health.

The aim of this study is to provide an overview of the impact of the COVID-19 pandemic and lockdown on the prevalence and clinical profile of suicide and SH in our ED. We compared the prevalence and clinical profile of SH in the ED during the COVID lockdown period in Nepal with the matching period of the previous year and the previous 3 months. We hope that all the stakeholders related to mental health will be primed to initiate mental health assessment and screening for the high-risk population and provide intervention for those needed during the period of crisis.

## METHODS

### Study design

This is a cross-sectional observational study of all consecutive fatal and nonfatal SH between March 24-June 23, 2020 (period of COVID lockdown-Period 1), matching periods in the previous year between March 24 and June 23, 2019 (Period 2) and 3 months period prior to the state of lockdown between December 24, 2019-March 23, 2020 (Period 3).

### Setting

The ED of Dhulikhel hospital-Kathmandu University Hospital (DH-KUH) which has approximately 20,000 visits annually and has a high acuity level as per internal audits. Most of the patients with acute SH present to the ED. The patients presenting to the ED are immediately triaged, categorized into different levels according to the severity and directed towards different designated areas. Ethical approval was obtained from the Institutional Review Committee, Kathmandu University School of Medical Sciences (IRC-KUSMS-69/20).

### Participants

Consecutive patients of all ages who presented to the ED during the study periods with any form of fatal or nonfatal SH including attempted hanging, impulsive self-poisoning, and superficial cuttings irrespective of the outcome were included. The patients who were discharged or referred were followed up with a telephone call to enquire about the patient’s final outcome. Incomplete data were excluded.

### Data Sources/measurement

The search in the electronic medical record (EMR) system with keywords suicide, attempted suicide, poisoning, hanging, self-harm, self-injury, overdose was conducted and the data was collected in the predesigned form. The final outcome was recorded combining the hospital records or phone calls if the case was referred, left against medical advice (LAMA), and discharged. The data from the EMR and phone calls were collected in a password secured laptop in the excel sheet.

### Quantitative variables

Variables studied include patient demographics (age, gender, address), mode of transportation, triage details, time to presentation in the ED, previous attempts of SH, past psychiatric illness, comorbidities, vital signs at presentation, investigations, treatment offered in the ED, duration of stay, and disposition of the patients. Disposition of the patients was divided into disposition from the ED and from the hospital. ED disposition was categorized into admission, discharge, LAMA, refer and mortality in the ED. Hospital disposition was categorized into discharge, LAMA, refer, and mortality in the hospital. The final outcome was categorized into recovery or mortality after follow-up phone calls and EMR review.

### Statistical methods

Data was analyzed with SPSS version 21. The categorical variables were expressed as frequency/proportion and continuous ones with mean with standard (SD) deviation or median with interquartile range (IQR) as appropriate. Categorical variables were compared with the chi-square test. The Independent-samples t-test/ANOVA or Man Whitney U test/Kruskal-Wallis test was used to compare the continuous variables with the categorical variables. *P*-value of less than 0.05 was considered significant.

## RESULTS

A total of 125 suicide/SH cases presented to the ED during the total study period, 55 during period 1 (44%), 38 during period 2 (30.4%) and, 32 during period 3 (25.6%) (Figure 1). The total number of patients presenting to the ED in all periods had decreased by 53% and 55.4% during the lockdown period when compared to previous periods (period 1:2085 versus period 2:3926 and period 3:3769 respectively). The cases of suicide and SH constituted 55 (2.6%), 38 (0.97%), and 32 (0.85%) among the total ED cases during periods 1, 2, and 3 respectively. Comparing the three periods, the cases of SH in period one increased by 44% (1.45 times) and 71.9 % (1.72 times) in relation to period two and three respectively.

**Figure 1:**
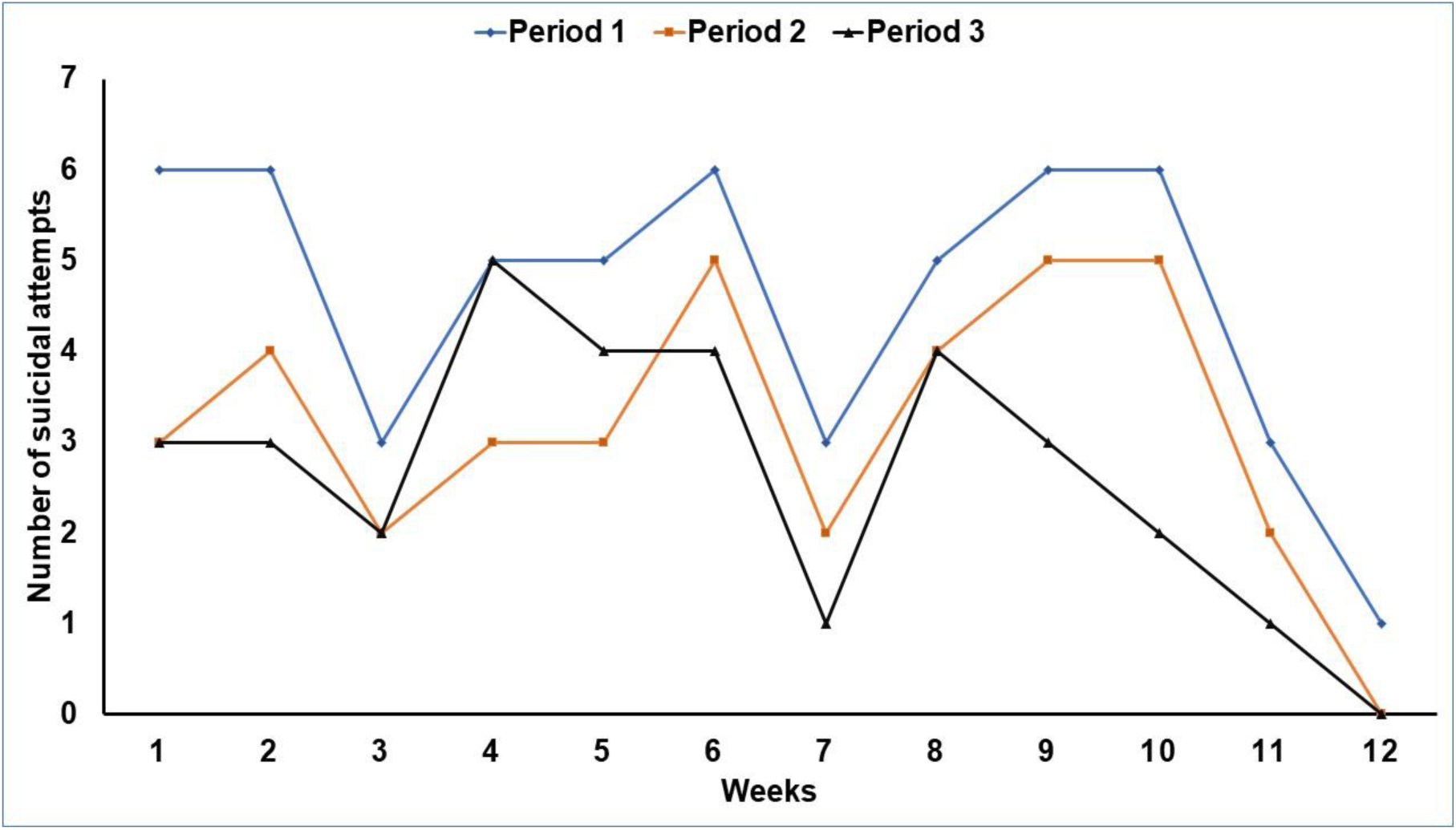
Number of visits to the ED with suicidal attempts throughout the 3 periods analyzed on a weekly basis. Period 1 (blue) (n=55): the lockdown period (March 24-June 23, 2020), period 2 (orange) (n=38): matching periods in the previous year (March 24-June 23,2019) and period 3 (black) (n=32): 3 months period prior to the state of lockdown (December 24 2019-March 23, 2020)

The comparison of different variables in relation to the three periods is depicted in table 1.

**Table 1:**
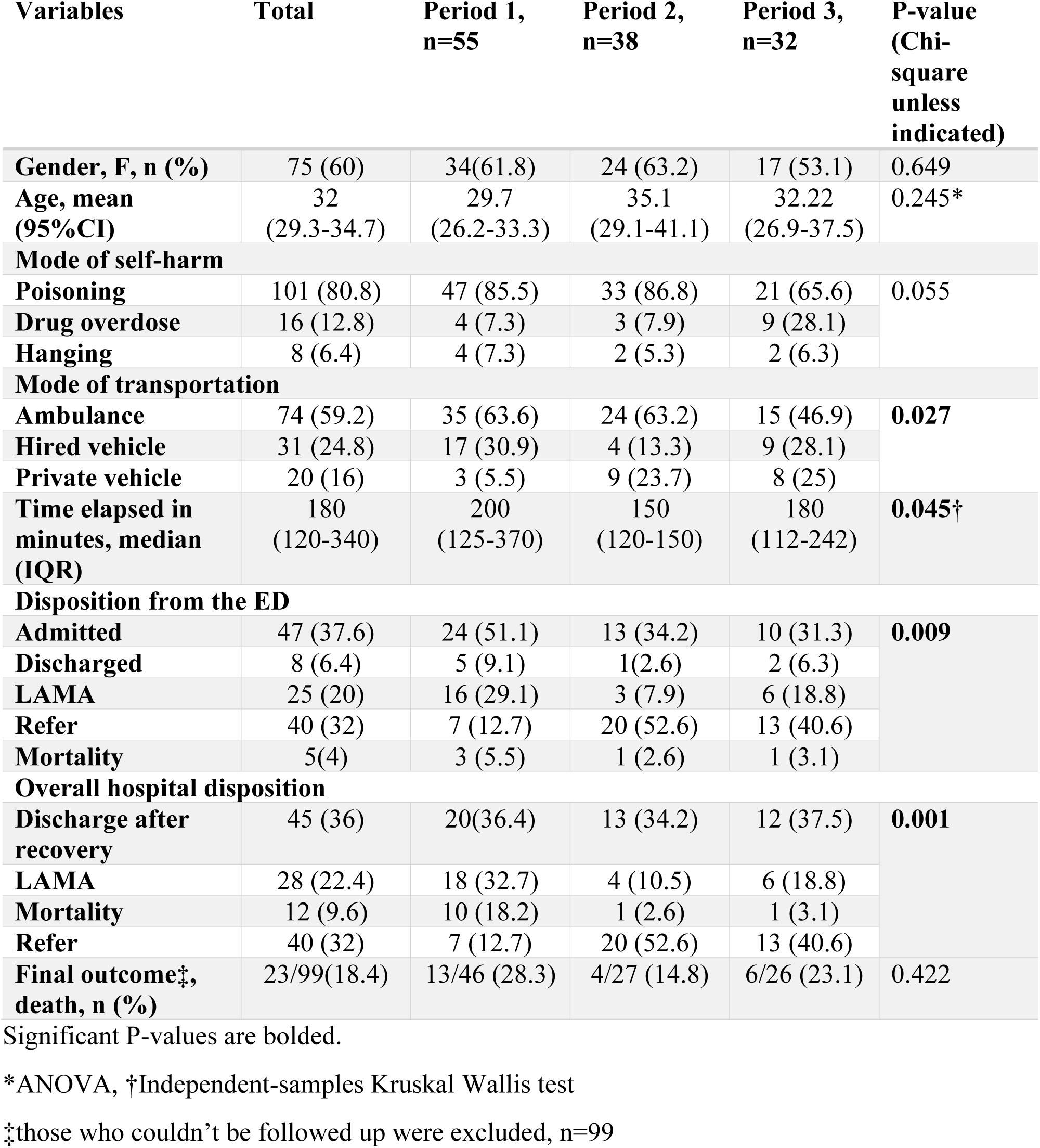
Different variables in relation to the 3 periods, n=125

The female gender was predominant 75 (60%) [period 1, 34(61.8%), period 2, 24 (63.2%) and period 3, 17 (53.1%)]. The gender difference was not statistically significant during all 3 periods (p-value=0.649). The mean age of the patients was 32 (95%CI: 29.3-34.7) {period 1 [29.7, 95% CI: 26.2-33.3], period 2 [35.1, 95% CI: 29.1-41.1], period 3 [32.22, 95% CI: 26.9-37.5]}. However, the difference was not significant (p-value = 0.245). Overall organophosphorus poisoning (OP) poisoning was the most common mode of suicidal attempt [66 (56.4%)], which was similar in all 3 periods [33 (64.7), 22 (61.1) and 11 (36.7) in periods 1, 2 and 3 respectively]. The dose of Atropine used ranged from 2 mg to 1660 mg [Median 57mg, IQR (20-200)]. More details on the mode of suicide and SH is illustrated in table 2. Multiple drugs or poison was ingested by eight patients (6.4%). Out of 117 cases of poisoning and drug overdose, 44 (37.6%) had induced vomiting at home. Sixteen patients (12.8%) required immediate airway management on arrival. Eight of them (6.4%) had an underlying psychiatric illness. Thirty-one (24.8%) patients were under influence of alcohol. The underlying reason for the suicidal attempt was not mentioned in the EMR for 53 (42.4%) patients. The alleged causes for suicidal attempts for the rest were disputes with family members, economic crisis, difficulty in coping with studies, disease or death of closed ones in 40(32%), 23 (18.4%), 4 (3.2%), and 5 (4%) cases respectively.

**Table 2:** Types of poison/drug overdose used for suicidal attempts, n=117

| Type of poison/drug overdose | n | % |
| --- | --- | --- |
| **Organophosphorus (OP)** | 66 | 56.4 |
| **Non-OP insecticide/fungicide** | 8 | 6.8 |
| **Aluminium Phosphide** | 5 | 4.3 |
| **Zinc Phosphide** | 3 | 2.6 |
| **Acetaminophen** | 10 | 8.5 |
| **Others (benzodiazepine, beta blockers, Ibuprofen, Alprazolam and others)** | 8 | 6.8 |
| **Unknown** | 11 | 9.4 |

There was a significant delay (p-value=0.045) in the arrival of the patients in period 1 [200 mins, IQR (125-370) vs 150 mins (120-150) and 180 (112-242) respectively in comparison to period 2 and 3]. There was a significant difference (p-value =0.009) in patient’s disposition from the ED, with an increase in the hospital admission rate and LAMA and a decrease in referrals. Similarly, the overall hospital outcome was also significantly different (p-value=.001), with increase in-hospital mortality (18.2% vs 2.6 % and 3.1%), and LAMA (18.2% vs 2.6% and 3.1%). Ninety-nine cases (79%) responded to a follow-up call which showed no statistical difference (p-value=0.422) in the overall mortality in the three periods (28.3%, 14.8%, and 23.1%).

## DISCUSSION

Despite a remarkable reduction in overall ED visits, our study showed a disproportionate increase in cases of suicide/SH during the lockdown period in comparison to the matching periods in the previous year and prior to the lockdown. OP poisoning was the most common mode of suicidal attempt during all periods. There was a delay in time to arrive at the hospital during the lockdown period with increased in-hospital mortality.

Suicide is a preventable loss that affects families, communities and entire countries. There is some evidence that deaths by suicide increased in Hong Kong during the 2003 SARS epidemic.[11] The WHO has predicted the rise in the number of mental health problems due to the global pandemic and has addressed this issue through various messages and publications related to mental health awareness and prevention.[18]. A recent report in China during COVID-19 revealed that about a third of their sample reported moderate to severe anxiety and 53 % of the respondents rate the overall psychological impact of the COVID-19 outbreak to be moderate to severe.[9] Strong restrictive measures to avoid COVID-19 infection have led to loneliness, loss of job, and loss of access to health which may precipitate or worsen the existing mental health problem. The lockdown has created a sudden economic recession, unemployment, worsened poverty which might have led individuals to contemplate suicide. Moreover, patients suffering from mental illnesses are unable to access health-care services. The effects might be worse in resource-limited countries like ours, where poor economic status is compounded by inadequate welfare support. Our study shows a considerable rise in the number of suicidal cases since the lockdown period. Various case reports of suicide-related to COVID-19 have been reported worldwide.[19] Studies from our neighboring countries, China,[12]India, [15] Pakistan,[13] and Bangladesh[14] have also raised concerns on increased suicide rate related to COVID-19. In contrast, a study exploring the mental health presentations in the ED before and during the COVID-19 outbreak in developed world showed decreased suicide and SH.[20,21] Gunnel et al have categorized COVID-19 related suicide risk factors into financial stressors, domestic violence, alcohol consumption, isolation, access to means, and irresponsible media reporting and published a public health response model to mitigate these risks. [22]) In our study, the common causes of suicidal attempts were disputes with the family members and economic crisis. No cases directly related to COVID-19 related illness or death were found.

Suicide is reported as the second leading cause of death among 15-29-year-olds globally. [4] Previous publications had reported higher suicide and SH among women, younger age group, migrant workers, the marginalized and disaster-affected population in Nepal. [5] After the 2003 SARS epidemics the increased suicidal rate among the elderly was reported in Hong Kong. [11] In our study, there was no statistically significant difference in the age of the patients attempting suicide in different periods. The mean age was 32 years and females attempted more suicide/SH in all the three periods. SH includes a variety of behaviors like hanging, self-poisoning, cutting, jumping from heights in response to intolerable mental pressure. OPs are the most commonly used form of pesticide in Nepal. [23] A previous study done at our ED showed that organophosphate poisoning was the commonest form of poisoning.[24]. Our study also showed that OP poisoning was the common cause of attempted suicide.

The lockdown, travel restrictions and social distancing likely contributed to a significant reduction in the use of private transport for transferal of patients to the ED. This may have caused the delay in presentation to the ED during the lockdown period in our sample. Our study also shows that the proportion of referrals of suicide/SH cases was less from our hospital during the lockdown period. During the lockdown period, the number of admissions for other illnesses requiring intensive care was low; therefore, more beds were vacant preventing referrals to other centers. This may be the reason for increased overall in-hospital mortality for suicide and SH during the lockdown period or they might have used lethal means to contemplate suicide.

The study was conducted in one of a rural tertiary care center, which is not representative of the whole country’s situation. Moreover, it doesn’t reflect the overall burden of mental health problems in the community and all population groups. An in-depth study of the cases was not done to determine the root cause of the increased suicidal attempts.

We found an increase in the number of patients presenting with suicide and SH in our ED during the pandemic which is likely to reflect an increased prevalence of mental illness in the community. To cope with the effects of the COVID-19 pandemic is emotionally challenging, especially for vulnerable individuals with underlying mental illness, low socio-economic status. Mental health problems are considered as a social stigma in our part of the world; therefore, people may be reluctant to share their feelings. Stressors like the increasing number of cases and deaths due to COVID-19, prolonged social isolation due to lockdown and social or physical distancing, economic regression and limited access to health care services due to fear of contracting COVID-19 may cause more panic, anxiety, and depression among the general public. The interplay of these factors in turn can precipitate suicide and SH in the future. Therefore, timely interventions to promote and protect the mental health of people and strategies to prevent suicide is of utmost importance.[25] Efforts to prevent the COVID-19 spread should be extended to raise awareness about dealing with mental health issues, recognizing warning signs of suicide and providing support to those needed. All the stakeholders, including policymakers, psychiatrists, psychologists, and other healthcare professionals should collaborate to raise awareness to screen, detect and timely intervene the needy patients. The challenge of the COVID-19 crisis might be an opportunity to advance the suicide prevention efforts in our country and thus to save many precious lives.

## Data Availability

Data will be available in special request

## Abbreviation

COVID-19: Coronavirus disease 2019
DH-KUH: Dhulikhel hospital-Kathmandu University Hospital
ED: Emergency Department
EMR: Electronic medical record
IRC-KUSMS: Institutional Review Committee, Kathmandu University School of Medical Sciences
LAMA: Leave against medical advice
OP: Organophosphate poisoning
SH: Self harm
SPSS: Statistical package for the social sciences
SARS: Severe acute respiratory syndrome
SD: Standard deviation
IQR: Interquartile range
WHO: World Health Organization

## Acknowledgements

ED staff of Dhulikhel Hospital-Kathmandu University hospital.

## Contributors

RS and APS conceptualized the study. SS1 and SS2 collected data. RS and APS analysed the data. RS wrote the initial manuscript draft, which was reviewed and developed with APS, SS1 and SS2. All authors reviewed and approved the final manuscript. RS takes responsibility for the paper as a whole.

## Funding Disclaimer

None

## Competing interests

None declared.

## Ethics approval

Ethical approval was provided by the institutional review committee of Kathmandu University School of Medical Sciences/Dhulikhel Hospital (KUSMS-DH, number: 69/20)

## Patient and public involvement

Patients or the public WERE NOT involved in the design, or conduct, or reporting, or dissemination plans of our research

## Notes

### Competing Interest Statement

The authors have declared no competing interest.

